# Generation-time distortion is an emergent property of epidemic control

**DOI:** 10.64898/2026.11.02.26367101

**Authors:** Kris V Parag, Patrick De Leenheer, Richard Pates

## Abstract

We find that a vast space of public health interventions necessarily reshape the intervals between successive infections, known as generation times, for directly-transmitted infectious diseases (e.g., COVID-19, Ebola virus, influenza). While some studies have connected specific interventions (e.g., contact-tracing, isolation) to shifted generation times, most epidemiological analyses assume these intervals are stationary. We reveal that this assumption is substantially more fragile than previously recognised. We develop a universal microscopic representation for all intervention mechanisms that reduce individual-level infectiousness and establish generation-time reshaping as a fundamental macroscopic feature of those interventions, emerging at the population level. By constraining the topology of our representation to reflect specific mechanisms, we explain and unify previous studies as well as predict generation-time phenomena yet to be observed. We further show that neglecting generation-time reshaping can under- or over-estimate intervention effectiveness and disease controllability by 100%, under realistic parameter settings. A stringent intervention can even produce no estimated transmissibility difference. Generation-time distortion causes unavoidable and underrated biases that can severely limit model-driven public health policy and pandemic preparedness.

## Introduction

Measuring the effectiveness of epidemic interventions is of fundamental public health importance. Prospective estimates of how available control actions limit transmission are critical for pandemic preparedness and have informed vaccination, lockdown and travel policies as well as biothreat risk assessments [1–4]. Retrospective evaluations of actions provide valuable evidence for critiquing decisions and have guided best practices on economic closures, mobility restrictions and contact tracing as well as formal enquiries into pandemic responses [5–8].

Most popular approaches for inferring effectiveness depend on the disease *reproduction number R* [9]. This is the average number of secondary infections caused by a primary infection and equals the epidemic branching parameter. An *R* above (below) 1 signals growing (declining) infections. Effectiveness *η* commonly involves comparing the pre-intervention *R* with its post-intervention value *R*_*k*_ [5, 6, 10]. Frequently, *η* := 1 − *R*_*k*_*R*^−1^ with a stabilising intervention achieving *R*_*k*_ ≤ 1 and *η* ≥ 1 − *R*^−1^. These formulae underlie how we assess epidemic controllability, severity, herd immunity and numerous pharmaceutical and non-pharmaceutical interventions [9–12]. However, they make a crucial assumption: that the random time intervals between primary and secondary infections are stationary [13]. These intervals comprise the *generation time distribution*, parametrised by *w*(*s*), the probability that this interval lasts *s* time units.

A handful of studies have questioned this assumption either finding mechanistic links between specific interventions such as contact tracing or isolation that modify generation times [14, 15] or uncovering empirical evidence of changes, again for specific interventions, from unusually high-resolution data on infection pairs [16, 17]. Although these works have highlighted the risk of biased estimates of *η*, they have revealed only modest biases over a small sample of possible interventions. For example, the most developed study in this space [14] presented a maximum relative bias of *θ* ≈ 18.5% (we define *θ* later).

Limited evidence of bias coupled with the costs of surveying infection pairs in real time, mean that the majority of policy-impactful research and intervention-effect estimates still assume stationary generation times [5, 6, 18]. We find this assumption to be unexpectedly fragile. We develop a Markov framework that provably represents any intervention that reduces infectiousness at the microscopic (individual) level and derive generation-time reshaping as an emergent macroscopic (population) level feature. We uncover universal biases in estimated effect that can cause *θ* = *±*100% under realistic settings. Commonly used estimators may even fail completely.

### Renewal processes and estimating effectiveness

We model an arbitrary epidemic as the macroscopic *renewal process* in Eq. (1) with *i*(*t*) counting new infections in our study location and *m*(*t*) counting imported infections. These counts average over microscopic transmission dynamics of individuals following a discrete-time Crump-Mode-Jagers branching process (see appendix).

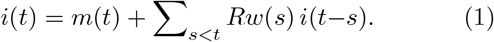

Prior to intervention, we assume the *w*(*s*) are known, *i*(*t*) and *m*(*t*) are observable and the population susceptible to infection is large so *R* is constant. The *infectiousness* of the infected cohort with age *s, i*(*t* − *s*), is *Rw*(*s*).

We unilaterally Z transform Eq. (1) to obtain the epidemic transfer function *G*(*z*) := *I*(*z*)*M* (*z*)^−1^ in Eq. (2) [19]. Here *z*^∗^ denotes the poles of *G*(*z*) and the dominant pole is the discrete-time *epidemic growth rate z*^∗^ = *r* [20].

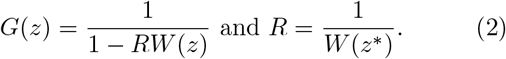

The *Euler-Lotka* relation in Eq. (2) links *R* with *r* [13]. Since ∑_*s*_ *w*(*s*) = 1, *W* (1) = 1. Critical epidemic stability (zero growth in infections) requires *R* = 1 and *r* = 1.

This renewal process (or its stochastic form) and the Euler-Lotka relation have been widely used to model out-breaks of COVID-19, SARS, Ebola virus, measles, influenza and many others [9, 21]. Under the standard assumption that *w*(*s*) is stationary, an intervention changes *r*, directly computable from the log gradient of *i*(*t*) [22], to *r*_*k*_. The estimated reproduction number resulting from Eq. (2), assuming a stationary *w*(*s*) is 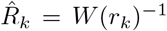. This intervention has estimated effect 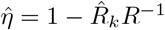.

This standard approach is our null model for intervention effectiveness. Our alternative model proposes that some *w*_*k*_(*s*) parametrises a post-intervention generation time distribution. This satisfies Eqs. (1) to (2) with subscript *k* replacements. The true intervention effect is then *η* in Eq. (3) and admits *θ*, the relative bias between estimating effects from the null and alternative models.

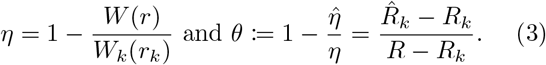

A positive *θ* indicates that we have under-estimated how much the intervention reduced transmission i.e., 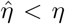. A negative *θ* means 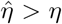. The extrema of relative bias are at *θ* = *±*1 and indicate that intervention effectiveness is under- or over-estimated by 100% of its actual value.

### Unified models of epidemic interventions

We formulate a universal but minimal microscopic intervention mechanism to test our effect models. Consider a newly infected individual so their age since infection is *s* = 0. On applying an intervention this individual is recurrently subject, as *s* ticks, to an arbitrary combination of detection and control stages (e.g., testing, antivirals, isolation) that suppress their ability to mix and transmit. While under control, the individual faces opportunities to exit (e.g., non-compliance, early release).

We can represent every such mechanism with an age-inhomogeneous *hidden Markov model* (HMM), in which the individual progresses through arbitrary mixing *M* (blue) and non-mixing *M*^*c*^ (red) states. We illustrate key mappings between primitive HMM motifs and interventions in Table I. Our representation covers all interventions that act by reducing infectiousness after infection. Motifs with non-mixing absorbing states model controls such as isolation and quarantine. Those with mixing absorbing states model release and non-compliance.

**TABLE I.**
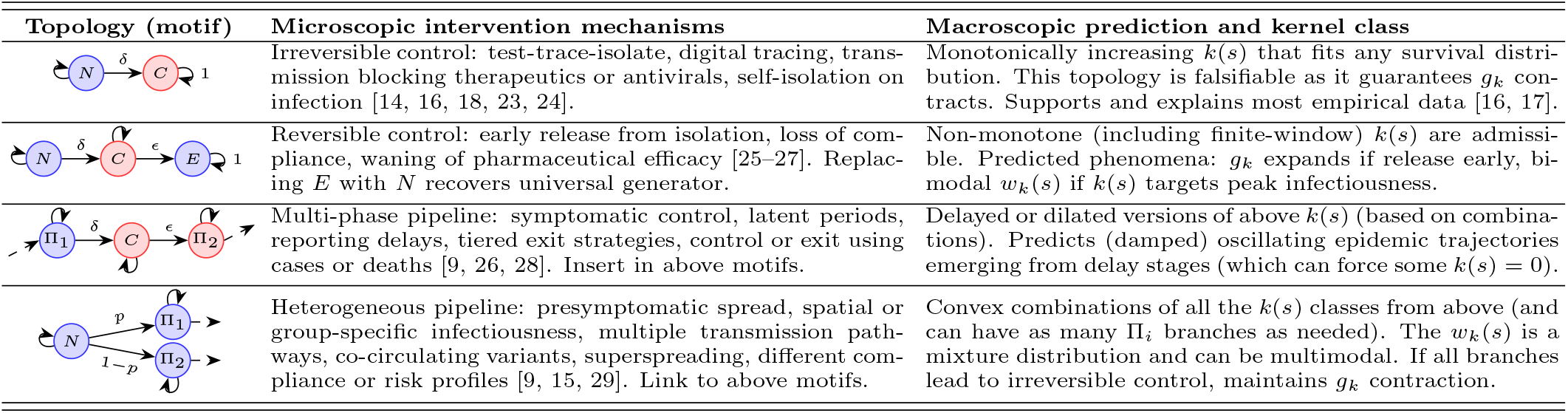
Microscopic motifs and macroscopic responses. We sketch topologies describing individual-level interventions. These constrain the population-level kernel classes (*k*(*s*)) that emerge and predict features of the post-intervention generation time distribution (*w*_*k*_(*s*), mean *g*_*k*_). States are normal (*N*), controlled (*C*) and exited (*E*) with generic detection (*δ*) and exit (*ϵ*) rates. The Π_*i*_ represent phase-type models that arbitrarily combine internal states and *p* is a splitting rate. Red states are non-mixing. These primitive motifs (or their combinations e.g., via dashed arrows) unify many epidemiological studies.

| Topology (motif) | Microscopic intervention mechanisms | Macroscopic prediction and kernel class |
|---|---|---|
| *N* δ *C* 1 | Irreversible control: test-trace-isolate, digital tracing, trans-mission blocking therapeutics or antivirals, self-isolation on infection [14, 16, 18, 23, 24]. | Monotonically increasing *k*(*s*) that fits any survival distri-bution. This topology is falsifiable as it guarantees *g*<sub>*k*</sub> con-tracts. Supports and explains most empirical data [16, 17]. |
| *N* δ *C* ϵ *E* 1 | Reversible control: early release from isolation, loss of com-pliance, waning of pharmaceutical efficacy [25–27]. Replac-ing *E* with *N* recovers universal generator. | Non-monotone (including finite-window) *k*(*s*) are admissi-ble. Predicted phenomena: *g*<sub>*k*</sub> expands if release early, bi-modal *w*<sub>*k*</sub>(*s*) if *k*(*s*) targets peak infectiousness. |
| Π<sub>1</sub> δ *C* ϵ Π<sub>2</sub> | Multi-phase pipeline: symptomatic control, latent periods, reporting delays, tiered exit strategies, control or exit using cases or deaths [9, 26, 28]. Insert in above motifs. | Delayed or dilated versions of above *k*(*s*) (based on combina-tions). Predicts (damped) oscillating epidemic trajectories emerging from delay stages (which can force some *k*(*s*) = 0). |
| *N* *p* Π<sub>1</sub> 1 − *p* Π<sub>2</sub> | Heterogeneous pipeline: presymptomatic spread, spatial or group-specific infectiousness, multiple transmission path-ways, co-circulating variants, superspreading, different com-pliance or risk profiles [9, 15, 29]. Link to above motifs. | Convex combinations of all the *k*(*s*) classes from above (and can have as many Π<sub>*i*</sub> branches as needed). The *w*<sub>*k*</sub>(*s*) is a mixture distribution and can be multimodal. If all branches lead to irreversible control, maintains *g*<sub>*k*</sub> contraction. |

We can extend both motifs using *phase-type* pipelines, which realise any positive-valued distribution by flexibly wiring internal states in series and parallel. These can model arbitrary delays to detection, treatment or release, presymptomatic spread, population heterogeneity, multiple transmission routes, risk-sensitive behaviour and targeted action, among others. By combining our primitive topologies, we can approximate arbitrarily complex intervention mechanisms. However, this description does not model interventions acting primarily through susceptibility reduction, such as shielding or vaccines and tracing that prevent infection instead of transmission.

In the appendix we prove that any microscopic HMM mechanism induces a macroscopic effect that is encoded in an age-dependent *kernel* 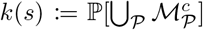. This is the emergent probability of not contributing to onward transmission at age *s*. The union ⋃ enumerates all HMM paths *P* to all non-mixing states up to times *s* 1. We further show that, when averaged across infected cohorts, we recover a post-intervention renewal process with the infectiousness of cohort *i*(*t* − *s*) reduced to *Rw*(*s*)[1 − *k*(*s*)]. Consequently, any intervention, regardless of complexity, once representable as an HMM induces a *k*(*s*) kernel. Distinct interventions with the same kernel have equal effect from a transmission perspective. We find a universal language for these kernels. Let *d*(*x*) and *e*(*x*) be the effective probabilities that an individual, infected *x* := *s* − 1 time units ago, is detected and controlled, or exits control. We construct a minimal HMM in Fig. 1 with states *M* = *N* (*x*), *M* ^*c*^ = *C*(*x*) and *k*(*s*) = P[*C*(*x*)]. For convenience we exclude age indices on HMM states.

**FIG. 1.**
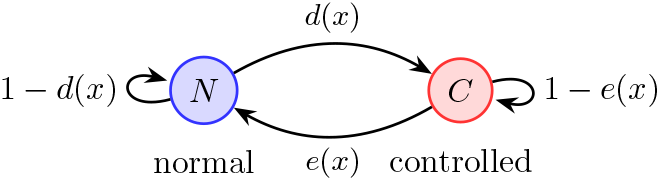
Universal intervention model. Normally mixing individuals with age *x* since their infection are detected and controlled with effective probability *d*(*x*). Control is exited or released with effective probability *e*(*x*). Mixing (non-mixing) states are in blue (red). The macroscopic effect on individuals at age *s* = *x*+1 is *k*(*s*), the probability of not occupying any mixing state at *x*. This HMM is a universal generator for all possible intervention kernels *k*(*s*) ∈ [0, 1] at all *s*.

This HMM admits Eq. (4) for how *d*(*s* − 1) and *e*(*s* − 1) contribute to *k*(*s*). Solving the recursion successively, we obtain Eq. (5), which enumerates over all the feedforward and feedback paths to state *C*(*x*) for ages up to *x*.

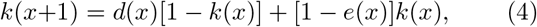

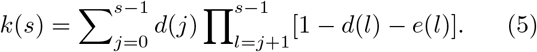

We prove Eq. (4) reaches every *k*(*s*) ∈ [0, 1]. Let Δ := *k*(*x*+1) − *k*(*x*) for any initial *k*(*x*). To realise any Δ *>* 0, we set *e*(*x*) = 0 and solve *d*(*x*)[1 − *k*(*x*)] = Δ. Larger *d*(*x*) increases *k*(*s*) and *d*(*x*) = 1 forces *k*(*s*) = 1. To realise any Δ *<* 0, we set *d*(*x*) = 0 and solve *e*(*x*)*k*(*x*) = −Δ. Larger *e*(*x*) decreases *k*(*s*) and *e*(*x*) = 1 forces *k*(*s*) = 0. To hold *k*(*s*) = *k*(*s* − 1), set *d*(*x*) = *e*(*x*) = 0. This establishes Fig. 1 as a universal generator of intervention kernels. Specific intervention topologies restrict the admissible kernel class as in Table I. Every infectiousness-reducing intervention is macroscopically equivalent to a 2-state HMM with some *d*(*x*) and *e*(*x*) probabilities.

### Generation time reshaping is fundamental

The macroscopic effects of infectiousness-reducing interventions are fully encoded by the kernel *k*(*s*). Applying control at time *τ*, we obtain (after some transient dynamics) the post-intervention renewal model of Eq. (6).

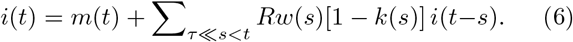

The true effect of the intervention and its threshold for stabilising the epidemic under any HMM mechanism follow as in Eq. (7). This generalises [9] and [19].

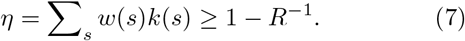

The epidemic is deemed controllable if the emergent *k*(*s*) of any deployable intervention fulfils this inequality. The controlled reproduction number is *R*_*k*_ = *R*(1 − *η*).

Importantly, the control kernel distorts the generation time distribution to Eq. (8), with two key consequences.

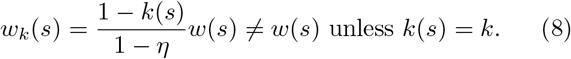

First, the prevalent stationary *w*(*s*) assumption is valid only if *k*(*s*) = *k* = *η* at all *s*. Mechanistically, this is implausible even for the universal generator in Fig. 1 as it requires the detailed balance condition of *d*(*x*)[1 − *k*] = *e*(*x*)*k* to hold over all *x*. Meeting this condition contradicts empirical data [26] because it implies that if our detection rate improves, our rate of releasing still infectious individuals from control must equally increase.

Many mechanisms from Table I restrict the *k*(*s*) class such that achieving a roughly fixed *k* becomes impossible. As examples, the irreversible motif has *k*(*s*) of Eq. (4) with *e*(*x*) = 0 and the multi-phase motif has *k*(*s*) = 0 for the *s* linked to delay stages. Neither admits any stationary solutions. The unchanged *w*(*s*) assumption is not emergent from microscopic dynamics.

Second, the universal reachability of *k*(*s*) means that generation time distortion is unavoidable and the level of distortion can be arbitrary. Any *w*_*k*_(*s*) absolutely continuous with respect to *w*(*s*) is realised by Fig. 1. Restrictions in Table I allow us to predict features of *w*_*k*_(*s*) that can corroborate or falsify hypotheses about intervention mechanisms. Defining the mean generation times *g* := ∑_*s*_ *sw*(*s*) and *g*_*k*_ := ∑_*s*_ *sw*_*k*_(*s*), we predict that when release rates are small (Δ *>* 0, *e*(*x*) small), *g*_*k*_ *< g* is certain. Generation time contraction is hence a macroscopic property of schemes like isolation.

This aligns with available data linking those interventions to changing generation times (or proxy serial intervals) [16, 17] and explains why faster detection (larger *d*(*x*) for small *x*) amplifies contraction. We make several predictions in Table I, to be tested when generation time surveillance improves. We posit that *g*_*k*_ *> g* expansion, a phenomena not yet observed [14], can emerge. Expansion is likely if interventions employ early detection (*d*(*x*) initially large, then small) but exit rates (*e*(*x*), Δ *<* 0) are large at low detection ages. We hypothesise this as a feature of premature release, fatigue or non-compliance.

### Intervention effect bias is universal and extreme

Pervasive and unmeasured intervention-induced generation time shifts imply ubiquitous bias when inferring intervention effect and epidemic controllability. We examine the extent of this bias under the standard stationary generation time assumption. We compute the controlled transfer function *G*_*k*_(*z*) = [1 − *R*_*k*_*W*_*k*_(*z*)]^−1^ with growth rate *r*_*k*_ using Eqs. (6) to (8). Relative bias *θ* follows from Eq. (3). Since *W* (*z*) ≠ *W*_*k*_(*z*), *θ* = 0 only when *r*_*k*_ = 1 as *W* (1) = *W*_*k*_(1) = 1. Consequently, except at critical stability (easily observable from a flat *i*(*t*) curve), bias is guaranteed and 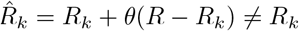.

As almost any *W*_*k*_(*z*) can emerge from an intervention, realisable biases may span− 1 ≤ *θ* ≤ 1. Hence we examine if the extrema of this range are realisable. Strikingly, both are achieved easily. The *θ* = −1 setting requires 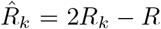 and *R <* 2*R*_*k*_ *<* 2 and overestimates the effect by 100% with 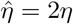. The *θ* = 1 setting is surprising as, it means that the intervention effect is completely nullified i.e., under-estimated by 100% with 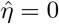.

This occurs when *RW* (*r*_*k*_) = 1 and yields 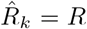. If *r*_*k*_ is a real subdominant pole of Eq. (2) and equally the dominant pole of *G*_*k*_(*z*), then 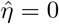. While this null effect is not deceptive if *R >* 1 and *r*_*k*_ *<* 1 as *i*(*t*) will visibly decay, it is problematic if *R <* 1 and we aim to regulate endemic infections [30]. Flaws in standard estimates of intervention effect can therefore be frequent and severe. In Fig. 2 we realise both extrema at plausible settings. Panel A models *w*(*s*) as a shifted geometric distribution with *g*(*r* − 1) = *R* 1 from Eq. (2). Panel B applies a shifted negative binomial *w*(*s*) with 2 poles satisfying 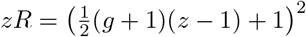. We use an irreversible *k*(*s*) kernel (see Table I) that implies a constant effective detection rate *d*(*x*). Values for *d*(*x*) and resulting contractions from *g* to *g*_*k*_ are consistent with data [16, 31]. We find that 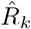 can be over 10 times smaller or larger than *R*_*k*_ and *g*_*k*_ *< g* does not inform on the sign of the bias.

**FIG. 2.**
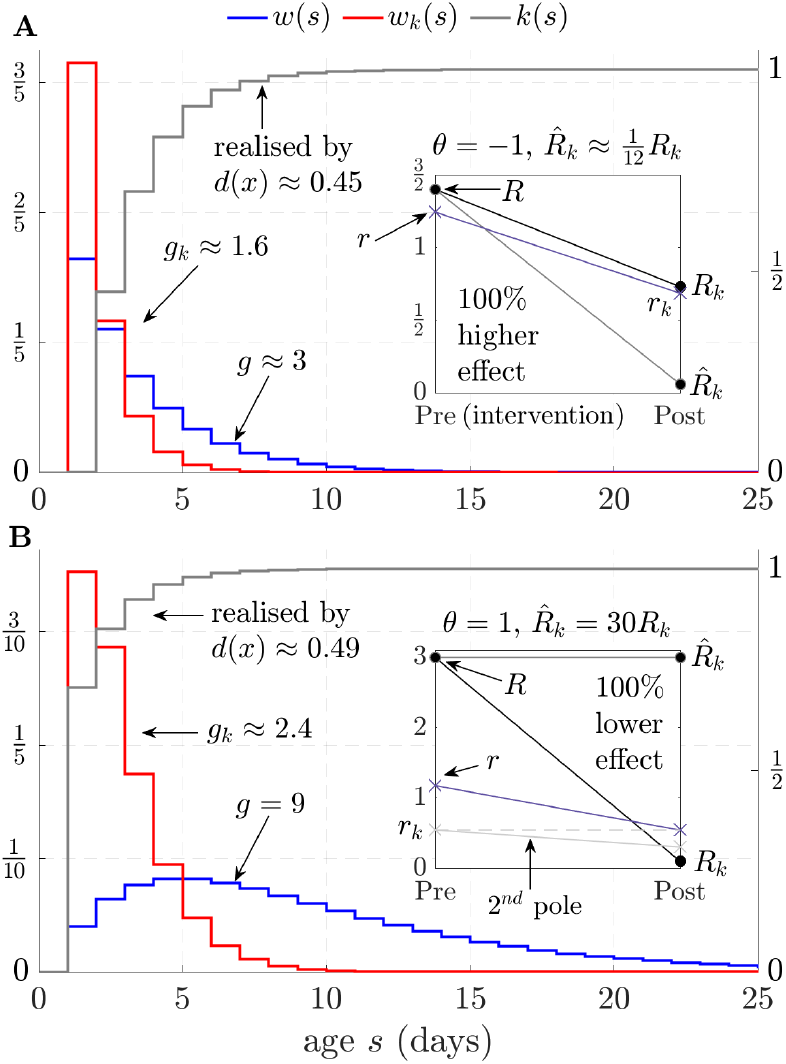
Extreme but plausible bias. We find practical interventions that completely devalue predominant approaches to estimating their effect. Removing infections according to *k*(*s*) shifts the disease generation time distribution from *w*(*s*) (with mean *g*) to *w*_*k*_(*s*) (mean *g*_*k*_), reproduction number from *R >* 1 to *R*_*k*_ *<* 1 and epidemic growth rate from *r >* 1 to *r*_*k*_ *<* 1. We know *R, w*(*s*) and measure *r*_*k*_. However, because we make the common assumption of inferring the effect of control using *w*(*s*), our estimate of *R*_*k*_ is 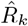. Panel A shows a case where we over-estimate the intervention effect by 100% and infer 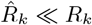 (relative bias *θ* = −1). Panel B realises a surprising scenario. We under-estimate the intervention effect by 100% (*θ* = 1), inferring 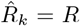 because the uncontrolled epidemic has a second pole matching *r*_*k*_. These extrema occur under irreversible control (Table I), fixed detection rate *d*(*x*) (Fig. 1) and the *g* to *g*_*k*_ contractions align with data [16].

### Transient but crucial intervention-driven effects

Because the breadth and depth of intervention-induced generation time shifts has been underrated, several consequent dynamical phenomena have been under-explored. We demonstrate in Fig. 3 how even interventions with the same true effect *η* (and hence equal *R*_*k*_) excite markedly different transient dynamics. We use COVID-19 parameters from [1] and compare interventions that reduce early (grey) and peak (red) infectiousness with the unrealistic *k*(*s*) = *k* action that preserves *w*(*s*) (blue). The grey and red actions achieve similar *r*_*k*_ and induce minimal bias under a stationary generation-time assumption.

**FIG. 3.**
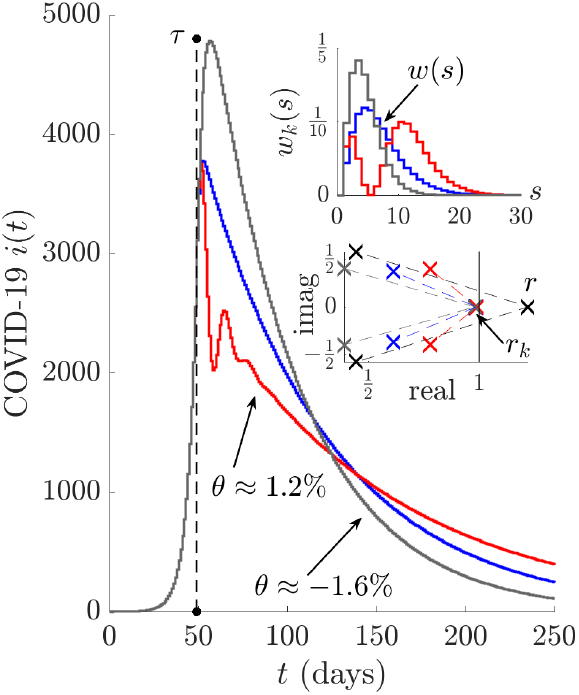
Subdominant pole effects. Intervention-induced generation time (*w*_*k*_(*s*), inset) changes relocate subdominant poles in epidemiologically important ways. We simulate COVID-19 epidemics pre- (times *t < τ*) and post-intervention with actions reducing early (grey) and peak (red) infectiousness and compare against the baseline of constant control, which maintains the generation time distribution *w*(*s*). All schemes have the same controlled reproduction number (*R*_*k*_), growth rate (*r*_*k*_) and small relative bias *θ* under stationary *w*(*s*) assumptions. Substantial incidence *i*(*t*) differences result from subdominant pole variations (x markers, inset).

Nonetheless, the differing *i*(*t*) curves in Fig. 3 are epidemiologically significant. The grey (irreversible) control mimics testing delay before isolation and causes a 26% larger peak. The red (reversible) action targets high transmissibility, sharply curbing infections, but has potential for oscillations and longer elimination times (it has the largest spectral gap, achieved by complex poles). Analyses assuming a fixed *w*(*s*) only excite the blue dynamics. Failing to recognise these second order, subdominant pole effects may cause such variations to be mis-attributed to other drivers such as behaviour or climate, further confounding evaluation of control decisions.

## Conclusion

We have established generation-time distortion as an emergent and universal property of interventions that reduce infectiousness. We found no mechanistic evidence to support the longstanding stationary *w*(*s*) convention in epidemiology and uncovered biases that are both significantly more extreme and common than previously known [14, 16]. Our results provide a principled route to refine intervention-effect estimation and controllability assessment, strengthening the rigour of public-health decision making and pandemic preparedness.

Our representation theorems reveal a descriptive hierarchy from intervention → topology → kernel → transfer function → poles that identifies what must be measured and what can be safely ignored. Neither resolving microscopic mechanisms nor imposing macroscopic simplifications are necessary to assess intervention effect. The sufficient object is the coarse-grained kernel *k*(*s*), which is measurable from pre- and post-intervention data and informs on effective control topologies. This supports calls for longitudinal infection-pair surveillance [32], converting this data into a targeted tool for policy comparison and mechanism identification.

Our analysis is limited to mean-field branching-process dynamics and interventions that suppress infectiousness. Network structure, susceptible depletion and time-varying policies (*k*(*s, t*)) may introduce further dynamical effects. Moreover, mechanisms with the same meanfield *k*(*s*) may differ in higher-order properties such as outbreak variability and extinction probability. Extending our coarse-graining framework beyond mean dynamics will establish when such microscopic details re-enter. By developing an exact coarse-graining and intervention-equivalence principle, this work provides a foundation for a needed general theory of epidemic interventions [33].

## Data Availability

This study uses no data. Code behind the figures are available upon request.

## ACKNOWLEDGMENTS

K.V.P is funded by a Senior AI+ fellowship at King’s College London and from the MRC Centre for Global Infectious Disease Analysis (Reference: MR/X020258/1), through the UK Medical Research Council.

## APPENDIX

## References

[1] N. M. Ferguson, D. Laydon, G. Nedjati-Gilani, N. Imai, K. Ainslie, M. Baguelin, et al., Impact of non-pharmaceutical interventions (NPIs) to reduce COVID-19 mortality and healthcare demand, Report 9 (Imperial College London, 2020).

[2] R. Sonabend, L. K. Whittles, N. Imai, P. N. Perez-Guzman, E. S. Knock, T. Rawson, et al., Non-pharmaceutical interventions, vaccination, and the SARS-CoV-2 delta variant in England: a mathematical modelling study, Lancet 398, 1825 (2021).

[3] T. Hollingsworth, N. Ferguson, and R. Anderson, Will travel restrictions control the international spread of pandemic influenza?, Nat. Med. 12, 497 (2006).

[4] M. E. Halloran, I. M. Longini, A. Nizam, and Y. Yang, Containing bioterrorist smallpox, Science 298, 1428 (2003).

[5] S. Flaxman, S. Mishra, A. Gandy, H. J. T. Unwin, T. A. Mellan, H. Coupland, et al., Estimating the effects of non-pharmaceutical interventions on COVID-19 in Europe, Nature 584, 257 (2020).

[6] N. Haug, L. Geyrhofer, A. Londei, E. Dervic, A. Desvars-Larrive, V. Loreto, et al., Ranking the effectiveness of worldwide COVID-19 government interventions, Nat. Hum. Behav. 4, 1303 (2020).

[7] M. Lipsitch, T. Cohen, B. Cooper, J. M. Robins, S. Ma, L. James, et al., Transmission dynamics and control of severe acute respiratory syndrome, Science 300, 1966 (2003).

[8] M. Walport, J. Aston, W. Barclay, I. Boyd, C. Donnelly, C. Dye, et al., COVID-19: Examining the Effectiveness of Non-Pharmaceutical Interventions, Report 9 (The Royal Society, 2023).

[9] C. Fraser, S. Riley, R. M. Anderson, and N. M. Ferguson, Factors that make an infectious disease outbreak controllable, Proc. Natl. Acad. Sci. USA 101, 6146 (2004).

[10] C. M. Peak, L. M. Childs, Y. H. Grad, and C. O. Buckee, Comparing nonpharmaceutical interventions for containing emerging epidemics, Proc. Natl. Acad. Sci. USA 114, 4023 (2017).

[11] C. P. Farrington, On vaccine efficacy and reproduction numbers, Math. Biosci. 185, 89 (2003).

[12] K. T. D. Eames and M. J. Keeling, Contact tracing and disease control, Proc. Roy. Soc. B 270, 2565 (2003).

[13] J. Wallinga and M. Lipsitch, How generation intervals shape the relationship between growth rates and reproductive numbers, Proc. Roy. Soc. B 274, 599 (2007).

[14] M. Favero, G. Scalia Tomba, and T. Britton, Modelling preventive measures and their effect on generation times in emerging epidemics, J. R. Soc. Interface 19, 20220128 (2022).

[15] J. Dushoff and S. W. Park, Speed and strength of an epidemic intervention, Proc. Roy. Soc. B 288, 20201556 (2021).

[16] S. T. Ali, L. Wang, E. H. Y. Lau, X.-K. Xu, Z. Du, Y. Wu, et al., Serial interval of SARS-CoV-2 was shortened over time by nonpharmaceutical interventions, Science 369, 1106 (2020).

[17] K. Sun, W. Wang, L. Gao, Y. Wang, K. Luo, L. Ren, et al., Transmission heterogeneities, kinetics, and controllability of SARS-CoV-2, Science 371, 10.1126/science.abe2424 (2021).

[18] L. Ferretti, C. Wymant, M. Kendall, L. Zhao, A. Nurtay, L. Abeler-Dörner, et al., Quantifying SARS-CoV-2 transmission suggests epidemic control with digital contact tracing, Science 368, eabb6936 (2020).

[19] K. V. Parag, How to measure the controllability of an infectious disease?, Phys. Rev. X 14, 031041 (2024).

[20] B. Boldin, O. Diekmann, and J. A. J. Metz, Population growth in discrete time: a renewal equation oriented survey, J. Differ. Equ. Appl 30, 1062 (2024).

[21] A. Cori, N. M. Ferguson, C. Fraser, and S. Cauchemez, A new framework and software to estimate time-varying reproduction numbers during epidemics, Am. J. Epidemiol. 178, 1505–(2013).

[22] K. V. Parag, R. N. Thompson, and C. A. Donnelly, Are epidemic growth rates more informative than reproduction numbers?, J. R. Stat. Soc. A 185, S5–S15 (2022).

[23] B. She, R. L. Smith, I. Pytlarz, S. Sundaram, and P. E. Paré, A framework for counterfactual analysis, strategy evaluation, and control of epidemics using reproduction number estimates, PLoS Computat. Biol. 20, e1012569 (2024).

[24] A. J. Black, T. House, M. J. Keeling, and J. V. Ross, Epidemiological consequences of household-based antiviral prophylaxis for pandemic influenza, J. R. Soc. Interface 10, 20121019 (2013).

[25] B. J. Quilty, S. Clifford, J. Hellewell, T. W. Russell, A. J. Kucharski, S. Flasche, et al., Quarantine and testing strategies in contact tracing for SARS-CoV-2: a modelling study., Lancet Public Health 6, e364 (2021).

[26] M. E. Kretzschmar, B. Ashby, E. Fearon, C. E. Overton, J. Panovska-Griffiths, L. Pellis, et al., Challenges for modelling interventions for future pandemics, Epidemics 38, 100546 (2022).

[27] P. Ashcroft, S. Lehtinen, D. C. Angst, N. Low, and S. Bonhoeffer, Quantifying the impact of quarantine duration on covid-19 transmission, eLife 10, e63704 (2021).

[28] K. V. Parag, C. Donnelly, and A. Zarebski, Quantifying the information in noisy epidemic curves, Nat. Comput. Sci 2, 584–(2022).

[29] T. Urmi, B. Pant, A. Dewey, I. Quintana-Mathe, I. Lang, J. Druckman, et al., Characterizing population-level changes in human behavior during the COVID-19 pandemic in the United States, Proc. Natl. Acad. Sci. USA 122, e2500655122 (2025).

[30] K. V. Parag and P. Hoscheit, Why do border controls fail to stabilise epidemics?, in 64th IEEE CDC (IEEE, 2025) pp. 5750–5755.

[31] G. Pullano, L. Di Domenico, C. E. Sabbatini, E. Valdano, C. Turbelin, C. Debin, et al., Underdetection of cases of COVID-19 in France threatens epidemic control, Nature 590, 134–(2021).

[32] M. U. G. Kraemer, O. G. Pybus, C. Fraser, S. Cauchemez, A. Rambaut, and B. J. Cowling, Monitoring key epidemiological parameters of SARS-CoV-2 transmission, Nat. Med. 27, 1854 (2021).

[33] A. Torneri, P. Libin, G. Scalia Tomba, C. Faes, J. G. Wood, and N. Hens, On realized serial and generation intervals given control measures: The COVID-19 pandemic case, PLOS Computat. Biol. 17, e1008892 (2021).

