## supplementary information for "Generation-time distortion is an emergent property of epidemic control"

### Supplemental Material for "Generation-time distortion is an emergent property of epidemic control"

Kris V Parag[1,2], Patrick De Leenheer[3], and Richard Pates[4]
[1]*Department of Engineering, King's College London, London, UK.*
[2]*MRC Centre for Global Infectious Disease Analysis, Imperial College London, London, UK.*
[3]*Department of Mathematics, Oregon State University, Corvallis, OR, USA.*
[4]*Department of Automatic Control LTH, Lund University, Lund, Sweden.*

#### I. DISEASE SPREAD

We model an epidemic as a discrete-time Crump-Mode-Jagers branching process [1]. An individual's age is the time since its infection. At each age, an infected individual is in one of finitely many states, and its infectiousness may depend on both. The state is a label for any other feature that matters, such as a stage of disease, a subpopulation or a stage of an intervention.

A branching process makes two simplifying assumptions. First, an individual inherits only its starting state, which fixes the distribution of its trajectory of states and infections. Given that state, the trajectory is independent of everything else in the epidemic except the individual's own descendants. Second, how an individual infects others at each age does not change with time or with how many are infected. Contact tracing breaks the first assumption, and susceptible depletion and seasonality break the second. These lie outside the model. We add one assumption of our own. An individual's current infectiousness depends on its past only through its current state, so a state's infectiousness does not depend on how it was reached. In return, an intervention can be modelled exactly as a rule for how individuals move between states, which leaves the infectiousness of every state unchanged.

We capture the above mathematically as follows. An individual is in state $X(s)$ at age $s$. It causes $Y_h(s)$ new infections starting in $h$ at that age. Its probability of being in state $h$ at age $s$, having started in $j$, is

$$(\boldsymbol{\Phi}(s))_{hj} = \mathbb{P}(X(s) = h \mid X(0) = j). \tag{1}$$

Its mean infections into $h$ at age $s$, when in state $j$, are

$$(\boldsymbol{W}(s))_{hj} = \mathbb{E}(Y_h(s) \mid X(s) = j). \tag{2}$$

The matrix $\boldsymbol{W}(s)$ is non-negative and bounded. It is the zero matrix at $s = 0$. At time $t$, $(\boldsymbol{i}(t))_h$ is the expected number of new infections starting in $h$. Its imported part is $(\boldsymbol{m}(t))_h$, which is finite. No one is infected before $t = 0$. Formally,

A1. $Y_h(s)$ depends on its past states only through $X(s)$,

$$\mathbb{E}(Y_h(s) \mid X(0), \ldots, X(s)) = (\boldsymbol{W}(s))_{hX(s)}. \tag{3}$$

Reference [1] states the backward form of the following equation for one ancestor without proof. We prove the forward form with imports and several starting states.

**Proposition 1.** *Under Assumption A1, expected incidence by starting state obeys*

$$\boldsymbol{i}(t) = \boldsymbol{m}(t) + \sum_{s \geq 1} \boldsymbol{W}(s)\boldsymbol{\Phi}(s)\boldsymbol{i}(t-s). \tag{4}$$

*Proof.* For an individual starting in $j$, column $j$ of $\boldsymbol{\Phi}(s)$ gives the distribution of its state at age $s$. By Assumption A1, averaging row $h$ of $\boldsymbol{W}(s)$ over it gives its mean infections into $h$ at age $s$, $(\boldsymbol{W}(s)\boldsymbol{\Phi}(s))_{hj}$. By the independence and identical distribution in Section I, those infected at time $t-s$ in state $j$ cause $(\boldsymbol{W}(s)\boldsymbol{\Phi}(s))_{hj}(\boldsymbol{i}(t-s))_j$ on average. No one infects at age zero or is infected before $t = 0$. Summing over $s \geq 1$ and $j$ and adding imports gives Eq. (4). Bounded $\boldsymbol{W}(s)$ and finite imports make each mean finite, by induction on $t$. □

#### II. INTERVENTION

In this section we show that, under two further assumptions, Proposition 1 reduces to the main paper's post-intervention renewal process. The assumptions simplify the model, but it can still capture detection, isolation, release and non-compliance at any age. States can still record disease stages or subpopulations that the intervention treats differently. Critically, the intervention then acts through one function $k(s)$, the probability of being in a non-mixing state at age $s$.

The first assumption concerns the intervention. It splits the states into mixing states, in which an individual infects others, and non-mixing states, such as quarantine, in which it infects no one and so does not branch. The second assumption concerns spread among mixing individuals. Each infects others alike on average, and each new infection starts in the same way, whoever caused it. Mixing states that differ in infectiousness break the first part, and groups that mix mainly among themselves break the second.

We capture this mathematically as follows. The mixing states form the set $\mathcal{M}$, and the non-mixing states, which cause no infections, the set $\mathcal{M}^c$. By the second assumption, every mixing state causes the same mean infections $\beta(s)$ at age $s$, and every local infection starts in a state drawn from one distribution $\boldsymbol{\pi}$, whoever the infector. Then $R = \sum_{s\geq 1}\beta(s)$, taken positive and finite, is the reproduction number of an individual that always mixes, and $w(s) = \beta(s)/R$ is its generation-time distribution, as in the main paper. Formally, with $\pi_h$ the probability of starting in $h$, the two assumptions are

A2. $(\boldsymbol{W}(s))_{hj} = \beta(s)\pi_h$ for $j$ in $\mathcal{M}$, and $(\boldsymbol{W}(s))_{hj} = 0$ for $j$ in $\mathcal{M}^c$.

The second assumption must also hold for imports. Each import starts in a state drawn from $\boldsymbol{\pi}$. Quarantine at the border breaks this. Formally,

A3. $\boldsymbol{m}(t) = m(t)\boldsymbol{\pi}$, with $m(t)$ the expected imports.

The following theorem reduces $\boldsymbol{\Phi}(s)$ to the kernel

$$k(s) = \mathbb{P}(X(s) \in \mathcal{M}^c), \tag{5}$$

with $X(0)$ drawn from $\boldsymbol{\pi}$. With the intervention acting from each infection and $m(0) = 0$, Eq. (6) is the main paper's Eq. (6). Reference [2] shows a continuous-time version for age-independent transitions.

**Theorem 1.** *Under Assumptions A1–A3, total expected incidence* $i(t) = \sum_h (\boldsymbol{i}(t))_h$ *obeys*

$$i(t) = m(t) + \sum_{s\geq 1} R\,w(s)(1 - k(s))i(t - s). \tag{6}$$

*Proof.* We show by induction on $t$ that $\boldsymbol{i}(t) = \boldsymbol{\pi} i(t)$, with $i(t)$ obeying Eq. (6). No one is infected before $t = 0$, so Eq. (4) and Assumption A3 give $(\boldsymbol{i}(0))_h = \pi_h m(0)$. By Eq. (5), $\mathbb{P}(X(s) \in \mathcal{M}) = 1 - k(s)$. Since $X(0)$ is drawn from $\boldsymbol{\pi}$, $(\boldsymbol{\Phi}(s)\boldsymbol{\pi})_j = \mathbb{P}(X(s) = j)$. If $\boldsymbol{i}(t - s) = \boldsymbol{\pi} i(t - s)$ for every $s \geq 1$, Eq. (4) and Assumption A2 give

$$\begin{aligned}(\boldsymbol{W}(s)\boldsymbol{\Phi}(s)\boldsymbol{\pi})_h &= \sum_{j\in\mathcal{M}} \beta(s)\pi_h \mathbb{P}(X(s) = j) \\ &= R\,w(s)\pi_h(1 - k(s)).\end{aligned} \tag{7}$$

By Eqs. (7) and (4), $(\boldsymbol{i}(t))_h = \pi_h(m(t) + \sum_{s\geq 1} R\,w(s)(1 - k(s))i(t - s))$, and summing over $h$ completes the induction. □

#### III. COMPUTING THE KERNEL

By Theorem 1, the intervention enters expected incidence only through $k(s)$. It follows that every model that satisfies Assumptions A1–A3 has a Markov chain on the same states with the same expected incidence. This chain is the main paper's hidden Markov model. Formally, let $(\boldsymbol{A}(s))_{hj}$ be the probability of moving from state $j$ at age $s$ to state $h$ at age $s+1$, and take column $j$ of $\boldsymbol{A}(s)$ to be the distribution of $X(s + 1)$ given $X(s) = j$, with $X(0)$ drawn from $\boldsymbol{\pi}$. Keeping $\boldsymbol{W}(s)$, $\boldsymbol{\pi}$ and the imports, the chain satisfies Assumptions A1–A3 with the same $k(s)$. It also satisfies the following assumption.

A4. $X(s)$ is a Markov chain with transition matrices $\boldsymbol{A}(s)$,

$$\mathbb{P}(X(s + 1) = h \mid X(0), \ldots, X(s)) = (\boldsymbol{A}(s))_{hX(s)}. \tag{8}$$

**Proposition 2.** *Under Assumptions A1–A4, for* $s \geq 1$, $k(s)$ *is the probability of the union of the paths of states that end in* $\mathcal{M}^c$*, as in the main paper, and*

$$k(s) = \sum_{h\in\mathcal{M}^c} (\boldsymbol{A}(s - 1)\cdots\boldsymbol{A}(0)\boldsymbol{\pi})_h. \tag{9}$$

*Proof.* By Eq. (5), $k(s)$ sums $\mathbb{P}(X(s) = h) = (\boldsymbol{\Phi}(s)\boldsymbol{\pi})_h$ over $h$ in $\mathcal{M}^c$. Summing Eq. (8) over the state at age $s$ gives $\boldsymbol{\Phi}(s + 1) = \boldsymbol{A}(s)\boldsymbol{\Phi}(s)$. With $\boldsymbol{\Phi}(0)$ the identity, $\boldsymbol{\Phi}(s) = \boldsymbol{A}(s - 1)\cdots\boldsymbol{A}(0)$. Expanding the product sums the probabilities of these paths, which are disjoint. □

#### IV. TWO-STATE MODEL

This section shows that, in expected incidence, the two-state model of the main paper's Fig. 1 realises every intervention that satisfies Assumptions A1–A3. Formally, the model has a mixing state $N$ and a non-mixing state $C$. At age $s$ an individual goes from $N$ to $C$ with probability $d(s)$, and from $C$ to $N$ with probability $e(s)$. In $N$ it causes $R\,w(s)$ new infections at age $s$ on average whatever its past, and in $C$ none. New infections and imports start in $N$, so the kernel, the probability of being in $C$, has $k(0) = 0$ and

$$k(s + 1) = d(s)(1 - k(s)) + (1 - e(s))k(s), \tag{10}$$

which is the main paper's Eq. (4).

**Theorem 2.** *Suppose a model satisfies Assumptions A1–A3 and has kernel* $k(s)$*. Then there are* $d(s), e(s) \in [0, 1]$ *for which the two-state model has kernel* $k(s)$ *for* $s \geq 1$*. With these and the same* $R$, $w(s)$ *and* $m(t)$*, both models have the same total expected incidence.*

*Proof.* Only $k(s)$ for $s \geq 1$ enters Eq. (6), so we take $k(0) = 0$. The construction after the main paper's Eq. (4) then gives $d(s), e(s) \in [0, 1]$ for which Eq. (10) yields $k(s)$ at every $s \geq 1$. The two-state model satisfies Assumptions A1–A3 with $\boldsymbol{\pi}$ on $N$, so by Theorem 1 both models obey Eq. (6) with the same $k(s)$, which fixes $i(t)$ from $m(t)$. □


[1] P. Jagers, General branching processes as Markov fields, Stoch. Process. Their Appl. **32**, 183 (1989).

[2] F. Avram, R. Adenane, D. Goreac, and A. Halanay, Explicit mathematical epidemiology results on age renewal kernels and $R_0$ formulas are often consequences of the rank one property of the next generation matrix, arXiv:2307.04774 (2023).
